# Postpartum continuity of care for women with HIV: Option B+ policy impact and limitations in South Africa

**DOI:** 10.1101/2025.07.19.25331807

**Authors:** Evelyn Lauren, Karl-Günter Technau, Kate Clouse, Nicola van Dongen, Amy Wise, Thalia Ferreira, Laura Rossouw, Jacob Bor, Mhairi Maskew

## Abstract

**Introduction:** Postpartum disengagement from HIV care is high, leading to adverse maternal outcomes and risk of transmission through breastfeeding. In January 2015, South Africa implemented Option B+, a policy extending lifelong antiretroviral therapy (ART) eligibility to all pregnant women living with HIV (PWLWH) regardless of CD4 count. We assessed the impact of Option B+ on postpartum continuity of care for PWLWH and identified predictors of disengagement in the Option B+ era.

**Methods:** We linked maternal data from Rahima Moosa Mother and Child Hospital in Johannesburg, South Africa, to HIV laboratory records from the National Health Laboratory Service. Using quasi-experimental designs (difference-in-differences, DID; regression-discontinuity, RD), we estimated the causal effect of Option B+ on postpartum retention in HIV care, defined as any HIV lab result 6-24 months after delivery. PWLWH with CD4 >500 cells/μL were newly eligible for lifelong ART under Option B+ (“treatment” group); PWLWH with CD4 ≤350 cells/μL served as “controls” as their eligibility for lifelong ART was unchanged by the policy. We also assessed predictors of postpartum disengagement from care in the Option B+ era.

**Results:** We included 1,684 women delivering 01/07/2013–30/06/2016, of which 14% delivered in the pre-Option B+ period. Option B+ was associated with a 19% increase (95% CI: 6%–33%) in postpartum retention for treatment group, relative to controls. We observed a 29% jump (95% CI: 7%–52%) in retention at the policy threshold in the treatment group. In the Option B+ era, PWLWH initiating ART during pregnancy had 76% lower odds of retention than those already on ART (OR: 0.24, 95% CI: 0.18–0.32).

**Conclusion:** Option B+ improved retention among PWLWH newly eligible for lifelong ART. Still, despite lifelong ART eligibility, women initiating ART during pregnancy exhibited significantly lower retention than those already on treatment, highlighting the need for further support to sustain care in this group.

**Key messages:** *What is already known on this topic:*

- Postpartum HIV care attrition is high among pregnant women living with HIV (PWLWH).
- The World Health Organization (WHO) published guidelines for preventing mother-to-child transmission of HIV, with Option B+ being the most recent guideline that extended lifelong ART eligibility to all PWLWH regardless of CD4 count.
- Existing research has reported suboptimal retention rates under Option B+, but has not specifically examined the impact on the population who gained eligibility under the new policy.

*What this study adds:*

- This study examined postpartum HIV care retention among PWLWH with high CD4 counts – the group newly eligible for lifelong ART under Option B+.
- We used quasi-experimental methods to estimate the causal effect of Option B+ by comparing PWLWH who delivered just before vs. just after the Option B+ policy was implemented, and we explored drivers of postpartum attrition from care.
- We demonstrated that Option B+ significantly increased HIV care retention in PWLWH with high CD4 counts; but that further efforts will be needed to support retention in care among PWLWH starting ART during pregnancy.

*How this study might affect research, practice, and policy:*

- Simplification of ART eligibility and expanded access to ART led to improvements in HIV care retention among PWLWH.
- However, PWLWH who initiated ART during pregnancy still experience high rates of attrition, highlighting the need for targeted interventions to support sustained care engagement for this group.

## Introduction

South Africa has made remarkable progress in reducing vertical transmission of HIV (VTP), with the national risk of six-week postpartum transmission plummeting from 25-30% before 2001 to approximately 1.4% in 2016 [1]. This success stems from the rapid and effective national scale-up [2,3] of the prevention of vertical transmission as part of the National Strategic Plan [4]. To achieve this, South Africa implemented a series of World Health Organization (WHO) recommendations specifically offering antiretroviral therapy (ART) to treatment-naïve pregnant women living with HIV (PWLWH) [5], before finally being replaced with the Universal Test-and-Treat (UTT) policy in September 2016 that made antiretroviral therapy (ART) available to all people living with HIV (PLHIV) regardless of CD4 count [6].

The WHO policies, named Option A, Option B, and Option B+, were implemented in April 2010 [7,8], April 2013 [9], and January 2015 [10] in South Africa, with each update providing further improvements and simplification of treatment regimens [5]. While all three policies provided lifelong ART for PWLWH entering care with CD4 <350 cells/μL, treatment guidelines differed for PWLWH with CD4 ≥350 cells/μL (see Table 1). Option A prescribed a complex multi-drug regimen that varied throughout pregnancy and delivery, while Option B simplified care by providing ART until breastfeeding cessation. Finally, Option B+ extended lifelong ART eligibility to all PWLWH regardless of CD4 count. These changes considerably reduced treatment barriers among PWLWH, with a particular benefit to treatment-naïve PWLWH with high CD4 counts.

**Table 1.** Baseline characteristics for the control (CD4 ≤350) vs. the treatment (CD4 >500) group, in the Pre-Option B+ vs. Post-Option B+ period (N=1,684)

| Baseline measure |  | Pre-Option B+ (N=240) |  | Post-Option B+ (N=1,444) |  |
| --- | --- | --- | --- | --- | --- |
|  |  | CD4 ≤350<br>(n=145) | CD4 >500<br>(n=95) | CD4 ≤350<br>(n=860) | CD4 >500<br>(n=584) |
| **Maternal age at delivery** | Median (IQR) | 29.9<br>(26.0, 34.1) | 26.2<br>(24.1, 30.9) | 30.7<br>(26.6, 35.2) | 30.8<br>(26.5, 34.8) |
| **Newly on ART** | %<br>(95% CI) | 85%<br>(79%, 91%) | 85%<br>(78%, 92%) | 82%<br>(80%, 85%) | 65%<br>(62%, 69%) |
| **CD4 count** | Median (IQR) | 210<br>(148, 279) | 639<br>(567, 768) | 233<br>(156, 294) | 640<br>(562, 770) |

Despite these policy improvements, postpartum adherence to HIV care remains a major challenge. Studies in South Africa consistently demonstrate that women who initiate ART during pregnancy face a heightened risk of disengagement from HIV care [11–14] compared to both men and non-pregnant women [11–13,15]. Poor maternal adherence to ART postpartum, in turn, can lead to new HIV infections transmitted through breastfeeding [16,17]. Postpartum retention rates in Sub-Saharan Africa remain suboptimal even under Option B+ [18–20], with a systematic review reporting retention rates below the general adult population [21]. However, prior studies have not specifically examined the effect of Option B+ on retention in care among treatment naïve PWLWH with high CD4 counts – the population that gained ART eligibility under the policy change. Our study leverages the introduction of Option B+ as a natural experiment to evaluate its causal impact on postpartum retention in HIV care and explored drivers behind care disengagement.

## Methods

### Data sources

We assessed the impact of Option B+ policy implementation on retention in HIV care among PWLWH in Johannesburg, South Africa, by linking the de-identified delivery data from the Rahima Moosa Mother and Child Hospital (RMMCH) maternal HIV cohort [22] to the laboratory care episodes (CD4/VL) from the National Health Laboratory Service (NHLS) HIV cohort [23] using a validated unique patient identifier.

From 2013 to 2020, RMMCH, with support from the Empilweni Services and Research Unit, collected data from women delivering at the hospital to monitor HIV-related birth care and track outcomes of HIV-exposed infants and their mothers [22]. The focus was on linking mothers and infants to support the prevention of vertical HIV transmission through maternal ART, infant prophylaxis, and early infant diagnosis. Initially focused on infant follow-up, data collection shifted to delivery and postnatal wards as infant birth HIV PCR testing became routine. Following low uptake of high-risk birth testing in 2013 and the introduction of universal birth testing in 2014, the cohort were expanded to capture comprehensive HIV care at delivery and build a full birth cohort of HIV-exposed infants.

NHLS is the sole provider of laboratory services for South Africa’s public sector health facilities, serving over 80% of the population [24]. Since 2004, NHLS maintains a centralized database of all laboratory records including HIV-related tests such as viral load and CD4 counts. Previously, a record linkage algorithm that combines probabilistic matching techniques with network analysis was used to construct the NHLS National HIV Cohort [cite 23, 24]. The linkage achieved high accuracy with a 1% overmatching and a 6% undermatching rate [23,25]. This cohort [23,25] enables longitudinal tracking of all lab-monitored patients enrolled in the public-sector HIV program, regardless of their movement between health facilities [26].

Unique patient identifiers used to link maternal records from RMMCH to the NHLS HIV cohort were obtained through direct stochastic matching of laboratory test barcodes [22]. Validation of a 10% manually matched sample indicated 71% of barcodes were exact matches, 22% matched on all variables except date of birth where date reversals are common, and 7% were unmatched.

### Study population

This analysis is nested within the RMMCH maternal HIV cohort. The cohort currently includes mothers known to be living with HIV or newly testing positive for HIV at delivery or thereafter who consented for data to be used for research purposes. From April to August 2013, enrolment occurred through counselling interviews. From September 2013 to May 2014, recruitment was expanded to include all high-risk babies qualifying for birth PCR testing. Since the implementation of universal birth testing in June 2014, recruitment was aimed at every exposed baby. Sporadic limitations in resources during 2018 and 2020 led to convenience sampling and somewhat lower coverage and enrolment rates observed in 2018 and 2020. All PWLWH enrolled in the RMMCH Maternal HIV cohort with delivery dates 18 months before and after the Option B+ policy went into effect in January 2015 were included in this analysis. Women giving birth between July 2013 and December 2014 were considered to be in the pre-period (Option B), while women giving birth between January 2015 and June 2016 were considered to be in the post-period (Option B+).

### Policy context

The Option B+ policy differs from the earlier Option B policy as follows:

Under Option B+, PWLWH with CD4 counts >350 cells/μL became eligible for lifelong ART. Under Option B, ART was dispensed only until cessation of breastfeeding. PWLWH with CD4 counts ≤350 cells/μL were eligible for lifelong ART under both Option B and Option B+. Coinciding with the implementation of Option B+, in January 2015, South Africa extended eligibility for ART to all people with CD4 counts from >350 to 500 cells/μL regardless of pregnancy [10].

### Treatment and control groups

PWLWH with CD4 >500 cells/μL became newly eligible for lifelong ART under Option B+ and are considered the “treated” population in this policy analysis; PWLWH with CD4 <350 cells/μL were eligible for lifelong ART in both Option B and Option B+ periods and are the “control” population; and PWLWH with CD4 350-500 gained eligibility under the separate global eligibility expansion and are excluded from the analysis.

Due to their new eligibility for lifetime ART under Option B+, we hypothesized that the policy change would increase longer-term retention in care among treatment naïve PWLWH with CD4 counts > 500 cells and have no effect on PWLWH with CD4 counts <350 cells.

### Outcomes

Our primary outcome was retention in HIV care 6-24 months postpartum, defined as the presence of an HIV laboratory test (CD4/VL) 6-24 months after delivery. In South Africa, PWLWH commonly discontinue breastfeeding within less than 6 months postpartum [27], after which, under Option B, PWLWH without clinical eligibility would discontinue ART. Thus, we hypothesized that the expanded lifelong ART eligibility introduced under Option B+ would primarily influence retention beyond 6 months postpartum.

The secondary outcome focused on understanding drivers of care attrition among PWLWH. Specifically, we examined whether PWLWH who initiate ART during pregnancy are less likely to be retained in care than those who initiated ART prior to pregnancy (selection effect), and whether birth event leads to postpartum attrition (causal effect).

### Exposures

Our primary exposure was policy era at the time of delivery. PWLWH who delivered on/after 1 January 2015 were classified exposed to Option B+, while those delivering before this date were considered unexposed (Option B). Given the possibility of delays in policy implementation, our results for this exposure follow an intention-to-treat (ITT) interpretation [28].

### Variable definitions ART history

PWLWH were classified into mutually exclusive groups based on their HIV care engagement prior to pregnancy. Individuals were classified as newly on ART during pregnancy if they were not in HIV care (had no VL) for at least 18 months before pregnancy. Otherwise, individuals were classified as continually on ART during pregnancy if they were engaged in HIV care (had any VL) for at least 18 months before pregnancy.

### CD4 count during pregnancy

PWLWH were classified into two groups based on their last CD4 count prior to delivery: ≤350 cells/μL and >500 cells/μL. PWLWH with CD4 >500 cells/μL gained lifelong ART eligibility under Option B+ and serve as the key exposed group. PWLWH with CD4 ≤350 cells/μL were eligible for lifelong ART in both policy periods and serve as our falsification group, as their eligibility status remained unchanged. We excluded PWLWH with CD4 counts between 351-500 cells/μL from our exposure groups as they became eligible for lifelong ART through separate clinical guideline changes in January 2015, regardless of pregnancy status [10].

### Statistical analyses

#### Impact of Option B+ policy on postpartum retention in care

We assessed the causal effect of the Option B+ policy on postpartum continuity in care through three separate quasi-experimental designs: differences-in-differences (DID), regression discontinuity (RD), and differences-in-discontinuities (DIDC).

We employed a DID approach to compare differences in postpartum retention over time between the treatment group (CD4 >500) and the control group (CD4 ≤350). We defined two policy periods: Option B (pre-intervention) and Option B+ (post-intervention). We modeled postpartum retention as a binary outcome using a standard linear regression model. Our model included indicators for treatment group, policy period, and an interaction term between the two. Our primary estimate of interest was the interaction term, which captures the difference in the change in retention attributable to Option B+ among PWLWH newly affected by the policy (CD4 >500). This estimate reflects the average treatment effect on the treated (ATT) [29]. We applied heteroskedasticity-robust standard errors to account for potential non-constant variance in the residuals.

A key assumption of the DID design is the parallel trends assumption, which states that, in the absence of the policy change, the difference in retention rates between the treatment and control groups would have remained constant over time. While our data include only two time periods, limiting our ability to examine pre-intervention trends, we assessed the plausibility of this assumption by comparing pre-policy baseline characteristics between the treatment and control groups, and additionally present DID models adjusting for baseline characteristics.

We used RD to compared postpartum retention among PWLWH with CD4 >500 cells/μL who delivered just before (control, Option B) vs. just after (treatment, Option B+) the guideline change using an RD design. For individuals near the threshold, the precise date of delivery was effectively random, making individuals on either side of the threshold exchangeable [30]. Assuming no manipulation to gain access to Option B+, women giving birth just before vs. after the policy change are as-good-as-randomly assigned to Option B vs. B+ and should be similar on both observed and unobserved factors.

We fit a standard linear regression model with an indicator for the policy period (Option B vs. Option B+), a continuous running variable (number of days from the delivery date to the policy change), and their interaction to allow for different slopes on either side of the threshold. We used a triangular kernel to assign greater weight to observations closer to the threshold, and applied heteroskedasticity-robust standard errors to account for potential non-constant variance in the residuals. Our model looks for evidence of change in intercept at the threshold, i.e., a jump in postpartum retention rates due to Option B+, while adjusting for potential differences in slope away from the threshold. This estimate reflects the local average treatment effect at the cutoff [31,32]. As a falsification test, we assessed for policy impacts among PWLWH with CD4 <350 cells/μL, whose eligibility did not change with Option B+.

The key assumptions of RD are exchangeability around the threshold and no manipulation of the running variable. To assess potential violations of these assumptions, we examined trends in baseline characteristics at the threshold (maternal age at delivery, ART history, CD4 count) to detect any compositional shifts that might indicate deviations from a randomized sample. We also conducted a McCrary density test to assess to detect any discontinuity in the running variable at the threshold, indicating potential manipulation.

Finally, we employed a DIDC design [33], comparing the change in retention rates at the policy threshold between PWLWH with CD4 >500 cells/μL (treatment group) and those with CD4 ≤350 cells/μL (control group). The DIDC design extends the RD design by differencing the discontinuities at the threshold across treatment groups, thus removing the possibility that other policies or factors occurring at the same time as Option B+ might confound the treatment effect estimate at the cutoff. We fit a standard linear regression model with an indicator for the policy period (Option B vs. Option B+), an indicator for the treatment group (CD4 >500 vs. ≤350), a continuous running variable (number of days from the delivery date to the policy change), and all 2-way and 3-way interactions. This specification allows for group-specific intercepts and slopes at either side of the threshold. We applied a triangular kernel weighting and heteroskedasticity-robust standard errors. Our estimate of interest is the interaction between the treatment group and the policy period, which corresponds to the difference in intercept shifts at the threshold between the CD4 strata. This DIDC estimate reflects the local average treatment effect on the treated at the cutoff.

We conducted sensitivity analyses using alternative model specifications to assess the causal effect of Option B+ on postpartum continuity of care. First, we adjusted for baseline maternal characteristics, including maternal age at delivery, ART history, and CD4 count as potential confounders in the quasi-experimental design models (DID, RD, DIDC). Second, we excluded observations from the first 3 months following policy implementation to allow for a transition period, ensuring that effects were estimated after the full implementation of Option B+.

#### Reasons for postpartum attrition in the Option B+ era

After assessing the causal effect of Option B+, we then investigated four hypotheses for the high rates of postpartum attrition observed among PWLWH relative to the general population:

##### Hypothesis 1

Attrition is high for new ART initiators regardless of pregnancy, and PWLWH are more likely than the general population to be new initiators (*new initiator effect*).

##### Hypothesis 2

Pregnant women starting ART because of their pregnancy rather than due to poor health (e.g. low CD4 count or clinical symptoms) may have weaker motivations for post-partum adherence (*healthy patient effect*).

##### Hypothesis 3

Pregnant women tend to be younger than the general population with HIV and younger PLHIV are less likely to be retained in care (*young age effect)*.

##### Hypothesis 4

The period encompassing pregnancy, childbirth, postpartum, and early infancy may present competing demands that could hinder maternal retention in care (*causal effect of childbirth*).

To test hypotheses 1 - 3, we fit a logistic regression model assessing the association of postpartum retention in care with prior ART experience (*new initiator effect*), CD4 count (*healthy patient effect*), and maternal age at delivery (*young age effect*). To test hypothesis 4, we conducted a matched cohort study comparing postpartum retention in care among ART-experienced women (i.e., women continually on ART) in our study sample and a matched sample of ART-experienced women from the general population in the NHLS laboratory database (controls). Each PWLWH in our sample was matched to three controls by test type (VL/CD4), number of years in HIV care (0, 1, ≥2), district (one of 52 in South Africa), age (±3 years), VL/CD4 test result (±25 units), and test date (±90 days). We then estimated the association between being in the childbirth sample and 6-24 month retention in care using a conditional logistic regression model accounting for the matched study design.

### Ethical considerations

Approval to analyze de-identified data was granted by the Institutional Review Board of Boston University (Protocol No. H-31968), the Human Research Ethics Committee of the University of the Witwatersrand (Protocol No. M200447) and NHLS Academic Affairs and Research Management System (Protocol No. PR2010539) with a waiver of informed consent.

The waiver was granted on the grounds that: (1) the study posed no more than minimal risk to participants; (2) the data had been previously collected as part of a laboratory database; (3) the data were de-identified; and (4) the research could not practicably be carried out without the waiver of consent due to the large number of persons in the database and because contacting these persons could introduce new risks including loss of privacy.

### Patient and public involvement in research

The study participants and the public were not involved in this study.

## Results

There were 1,684 PWLWH enrolled in the RMMCH Maternal HIV cohort who delivered between July 2013 and June 2016. Of these, 240 (14%) PWLWH delivered in the pre-Option B+ period, and 1,444 (86%) delivered in the post-Option B+ period (**Table 1**). Pre-policy baseline characteristics, including maternal age at delivery and proportion newly on ART, were comparable between the treatment (CD4 >500) and the control (CD4 ≤350) group, supporting the parallel trends assumption. Among PWLWH with CD4 >500 cells/μL, the proportion of PWLWH newly on ART were lower in the post-period (65%; 95% CI: 62%, 69%) compared to the pre-period (85%; 95% CI: 78%, 92%). McCrary density test showed no evidence of manipulation at the policy threshold (**Figure S1**). There were no significant differences in baseline characteristics, including maternal age at delivery, proportion newly on ART, and CD4 count, for PWLWH with CD4 ≤350 cells/μL at the policy threshold (**Table S1**). However, maternal age at delivery was significantly lower below the cutoff vs. above the cutoff for PWLWH with CD4 >500 cells/μL (difference: 3.5; 95% CI: 0.7, 6.3).

Retention at 6-24 months improved following the implementation of Option B+ among PWLWH with CD4 count >500 cells/μL. In this group, retention increased from 42% (95% CI: 32%, 52%) under Option B to 67% (95% CI: 64%, 71%) under Option B+ (**Table 2**). Among PWLWH with CD4 ≤350 cells/μL, retention rates increased from 58% (95% CI: 50%, 66%) to 64% (95% CI: 61%, 67%) with exposure to Option B+. Using a DID approach, we estimated that exposure to Option B+ was associated with a 19% increase (95% CI: 6%, 33%) in probability of postpartum retention for PWLWH with CD4 >500 cells/μL, compared to what would have been expected based on the control group’s (CD4 ≤350) trend (**Figure 1**). The DID estimates were robust to adjustment for differences in baseline characteristics (β=14%; 95% CI: 1%, 28%) (**Table 2**), and after excluding deliveries from January to March 2015 to allow for a transition period for the full implementation of Option B+ (**Table S2**).

**Table 2.** Causal effect of Option B+ on postpartum retention (N=1,684)

| A. Difference in Differences (DID) | Averages during each era |  | DID estimate (crude) |  | DID estimate (adjusted)* |  |
| --- | --- | --- | --- | --- | --- | --- |
|  | Option B | Option B+ | Pre/Post Difference | P-value | Pre/Post Difference | P-value |
| Target pop: CD4>500 | 42%<br>(32%, 52%) | 67%<br>(64%, 71%) | 19%<br>(6%, 33%) | 0.006 | 14%<br>(1%, 28%) | 0.041 |
| Control pop: CD4≤350 | 58%<br>(50%, 66%) | 64%<br>(61%, 67%) |  |  |  |  |
| B. Regression Discontinuity (RD) | Predictions just before/after Option B+ policy change |  | RD estimate (crude) |  | RD estimate (adjusted)* |  |
|  | Just before Option B+ | Just after Option B+ | Pre/Post Difference | P-value | Pre/Post Difference | P-value |
| Target pop: CD4>500 | 41%<br>(22%, 61%) | 71%<br>(60%, 81%) | 29%<br>(7%, 52%) | 0.011 | 22%<br>(2%, 43%) | 0.031 |
| Control pop: CD4≤350 | 56%<br>(41%, 72%) | 61%<br>(51%, 70%) | 4%<br>(-14%, 23%) | 0.640 | 4%<br>(-13%, 22%) | 0.639 |
|  | Difference in RD Estimates |  | 25%<br>(-4%, 54%) | 0.092 | 18%<br>(-8%, 45%) | 0.181 |

\*Estimates adjusted for maternal age at delivery (<26, 26-34, >34), ART history (newly on ART vs. continually on ART), and CD4 count value

**Figure 1.**
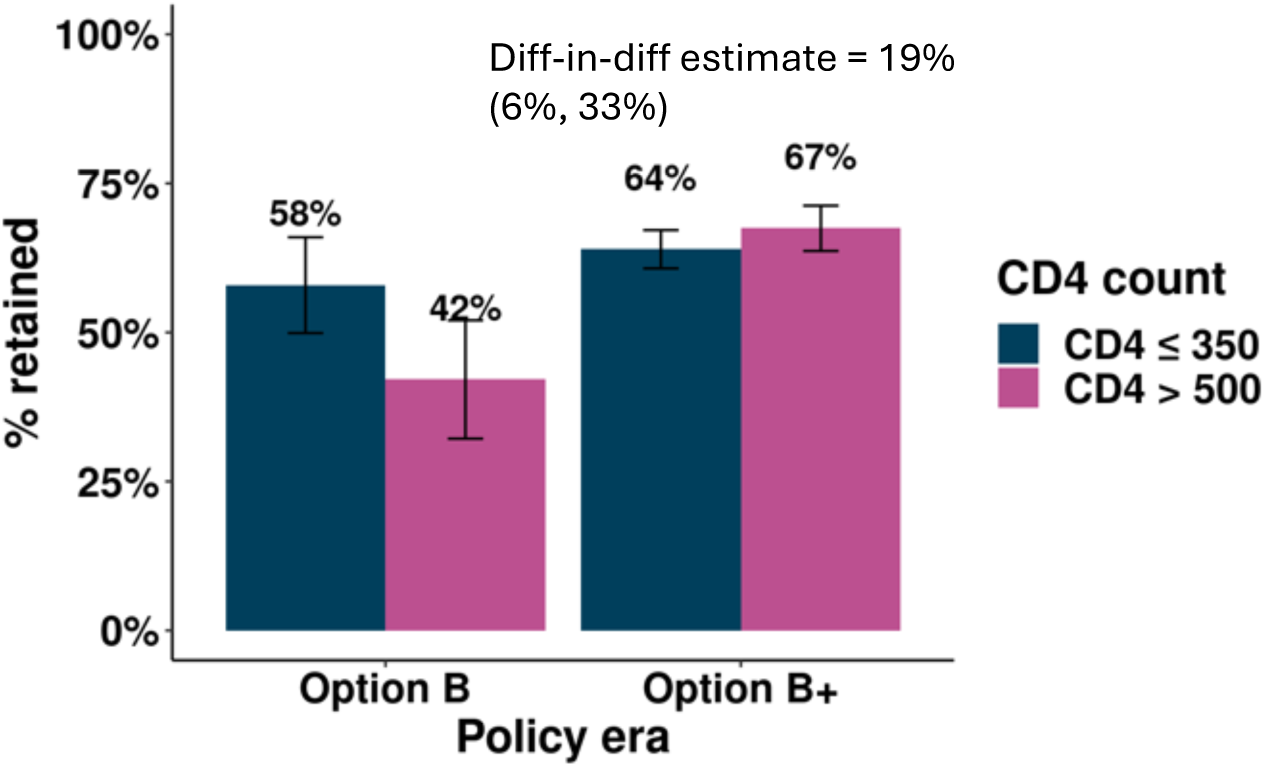
Differences-in-differences estimates for 6-24 months retention in care by CD4 count and policy era (N=1,684)

Using an RD design, we observed a 29% jump (95% CI: 7%, 52%) in 6-24 months retention around the policy threshold among PWLWH with CD4 >500 cells/μL, from 41% (95% CI: 22%, 61%) just below the threshold to 71% (95% CI: 60%, 81%) just above the threshold (**Figure 2, Table 2**). The RD estimates remained robust after adjusting for differences in baseline characteristics (β=22%; 95% CI: 2%, 43%) and excluding deliveries from January to March 2015 (**Table S2**). As expected, we observed no significant change in retention among PWLWH with CD4 count ≤350 cells/μL, whose eligibility did not change with Option B+ (**Table 2**). The DIDC estimate indicates a 25% greater increase in postpartum retention at the policy threshold among PWLWH with CD4 >500 cells/μL vs. those with CD4 ≤350 cells/μL, however the confidence interval on this estimate was wide (95% CI: -4%, 54%) (**Table 2**).

**Figure 2.**
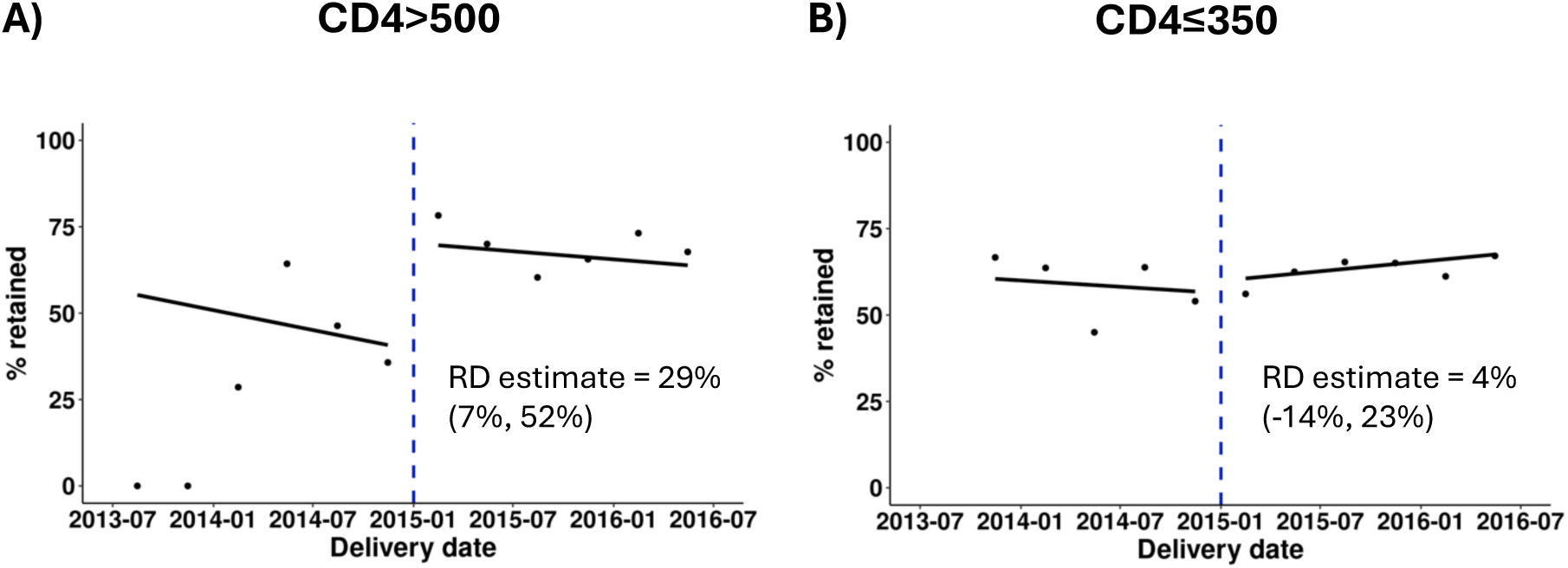
Regression discontinuity estimates for 6-24 months retention in care, with delivery date as running variable: A) For the target population (CD4 >500); B) For the control population (CD4≤350) (N=1,684)

Taken together, the results of our quasi-experimental policy analyses indicate that Option B+ increased post-partum retention in care by about 20 percentage points, with effects limited to PWLWH who were newly eligible for lifetime ART under the policy. Nevertheless, even in the Option B+ era, one-third of PWLWH were not retained on ART after childbirth. We now assess the reasons for high attrition among PWLWH.

In the Option B+ era, PWLWH newly on ART during pregnancy had 76% lower odds of being retained in care at 6-24 months postpartum compared to those who were already on ART prior to pregnancy (OR: 0.24, 95% CI: 0.18, 0.32), after adjusting for maternal age at delivery and CD4 count (**Table 3**). These findings provide a strong evidence of a “new initiator effect”: women initiating ART during pregnancy were significantly less likely to remain in care compared to those who were already on ART before pregnancy. Higher attrition from care amongst new initiators persisted even though these women were eligible for lifetime ART under Option B+.

**Table 3.** Predictors of postpartum retention in the Option B+ era (N=1,946)

| Characteristic | Odds ratio (95% CI) | P-value |
| --- | --- | --- |
| ART experience (ref: ART-experienced at conception) |  |  |
| Started ART during pregnancy | 0.24 (0.18, 0.32) | <0.001 |
| Maternal age at delivery (ref: <26 years) |  |  |
| 26-34 years | 1.34 (1.05, 1.71) | 0.017 |
| ≥35 years | 1.61 (1.21, 2.14) | 0.001 |
| CD4 count (ref: ≤200 cells/μL) |  |  |
| 201-350 cells/μL | 0.96 (0.72, 1.29) | 0.804 |
| 351-500 cells/μL | 0.85 (0.62, 1.16) | 0.306 |
| >500 cells/μL | 0.95 (0.71, 1.28) | 0.734 |

On the other hand, we did not find evidence of a “healthy patient effect”, as we found no significant gradient between CD4 count during pregnancy and retention in care. We did find significant associations between older maternal age and postpartum retention, highlighting excess risk of attrition among younger women.

Results from the matched cohort analysis showed that ART-experienced birthing women in our sample had 68% higher odds of being retained at 6-24 months compared with matched (ART-experienced) controls (OR: 1.68; 95% CI: 1.21, 2.34) (**Table S3**). These findings of higher retention among birthing women suggest that significant life-changing (and potentially life-disrupting) experiences of childbirth and parenting an infant do not in and of themselves cause women to exit HIV care after birth. Rather, our results suggest that recent mothers are more likely to be retained in care than the general population. Our analyses suggest that low rates of postpartum retention in care are primarily due to the high prevalence of new initiators among pregnant women and their relatively young age and are not a direct consequence of childbirth.

## Discussion

In our study at a maternity hospital in Johannesburg, South Africa, we found improved retention in HIV care in the Option B+ era among PWLWH with CD4 >500 cells/μL – the primary demographic affected by this policy change.

Prior to Option B+, we observed lower 6-24 months retention among PWLWH with CD4 >500 cells/μL, who were no longer eligible for ART after breastfeeding, relative to PWLWH with CD4<350 cells/μL who remained eligible for life. These results are consistent with previous findings which reported poor retention rates among individuals not eligible for ART [34–38], particularly among pregnant women [39]. Option B+, which extended lifelong ART eligibility to all PWLWH regardless of CD4 count, nearly equalized retention rates between those initiating ART with CD4 >500 cells/μL and those with CD4 ≤350 cells/μL. This finding aligns with previous studies which have shown that universal ART eligibility leads to improved retention rates [40–43].

Although prior research specifically focused on PWLWH who gained lifelong ART eligibility under Option B+ are limited, some have examined the association between CD4 count at initiation on care retention in the Option B+ era. These studies suggest that individuals initiating ART with higher CD4 counts have retention rates that are comparable to, or better than, those with lower CD4 counts. Specifically, a study in Cameroon found no significant association between CD4 count at initiation (≤350 vs. >350 cells/μL) and treatment discontinuation [44], while a study in Uganda reported higher retention rates among PWLWH with CD4 >350 cells/μL vs. ≤350 cells/μL at initiation [45].

Despite improved retention rates under Option B+, PWLWH who initiated ART during pregnancy continued to have significantly lower retention rates than those already on ART before pregnancy, consistent with findings from prior studies in sub-Saharan Africa [46–49]. Similar patterns have also been reported under Option A, Option B, and Option B+ [14,50,51], with low retention rates among PWLWH newly initiating ART, regardless of WHO recommendation. Lower maternal age was also significantly associated with lower retention, consistent with previous evidence [52,53]. Additionally, prior research has documented substantial loss-to-follow-up (LTFU) among pregnant women irrespective of baseline CD4 count [54], highlighting the need for targeted interventions beyond Option B+ to improve long term retention. We examined whether there was a causal effect of childbirth leading to lower retention, but found that retention was higher among ART-experienced postpartum women relative to matched controls in the general population.

Our study has several strengths. First, we have access to data from both the Option B and Option B+ periods and CD4 count information, allowing us to directly compare retention rates across these two policy periods among PWLWH whose treatment eligibility changed with this policy vs. those whose eligibility remained the same. We were able to observe women who delivered just before and immediately after the policy change and account for CD4 count and differences in baseline characteristics, enabling us to better isolate the effect of Option B+ on retention outcomes. Second, the linkage between the RMMCH and the NHLS laboratory database allows for comprehensive follow-up of women who continue receiving care within the public sector and provided access to HIV care history before pregnancy and entry to RMMCH. Women who switched facilities but remain in the public sector will still have their laboratory data recorded in the NHLS database, minimizing issues with LTFU.

Our study has several limitations. First, the sample size in our data, particularly during the Option B period, is low, resulting in reduced statistical power, lower precision, and increased sensitivity to noise. Therefore, these findings should be interpreted with caution and may not be generalizable to the broader population. Second, there is an imbalance in sample size between the Option B and Option B+ periods and limited data on confounders (e.g., socioeconomic status). However, this is unlikely to substantially impact our findings as the baseline characteristics of women in both periods are largely similar, and we accounted for differences in the baseline characteristics in our robustness checks. Third, due to limited data availability before 2013, we could not evaluate the impact of Option A and the initial Option B implementation period. Fourth, women who transition out of the public sector will be recorded as “not retained in care”, as their lab results are not captured in the NHLS database. Thus, our findings likely provide a conservative estimate of retention. Regardless, we anticipate the number of women transitioning out of the public sector to be relatively small as over 80% of the South African population access care within the public sector [55]. Fifth, our database lacks specific indicators to differentiate between treatment groups (Option B vs. Option B+, ART until breastfeeding cessation vs. lifelong ART). Therefore, we relied on proxy measures, such as delivery dates and CD4 counts before delivery, to assign treatment categories. This may have led to misclassification of some women. Sixth, the findings of this study are drawn from a single urban maternity hospital (RMMCH) which may limit their generalizability, particularly to rural or per-urban contexts. RMMCH serves a unique patient population, with a relatively high proportion of non-South African patients seeking care (39%) [22]. However, the proportion of patients receiving ART at RMMCH is comparable to the national average [22,56]. Lastly, under Option B+, CD4 testing was no longer required to determine immediate eligibility for ART. This simplification of eligibility criteria may have influenced care retention independently of the expanded eligibility of lifelong ART. However, findings from the RD analysis suggest that this pathway had minimal impact on retention.

## Conclusion

Option B+ resulted in improvements in retention rates for PWLWH newly eligible for lifelong ART under the policy. However, for many PWLWH, eligibility was not enough. Despite gains under Option B+, PWLWH who initiated ART during pregnancy still exhibited high postpartum attrition, with potential adverse impacts for their own health and HIV transmission through breastfeeding or sex. Low rates of retention among PWLWH relative to the general population are due primarily to the large number of PWLWH who start ART during pregnancy. Focused interventions are needed to improve retention in care within this group for improved maternal health and so support the elimination of vertical transmission.

## Data Availability

Data from the National Health Laboratory Service (NHLS) National HIV Cohort can be accessed through application to the NHLS Academic Affairs and Research Management System: aarms.nhls.ac.za.

