## Supplemental Tables and Figures for "Postpartum continuity of care for women with HIV: Option B+ policy impact and limitations in South Africa"

**Appendix**

**Figure S1.** McCrary density test to assess discontinuity in the number of deliveries at the threshold: A) For the target population (CD4 >500); B) For the control population (CD4≤350)

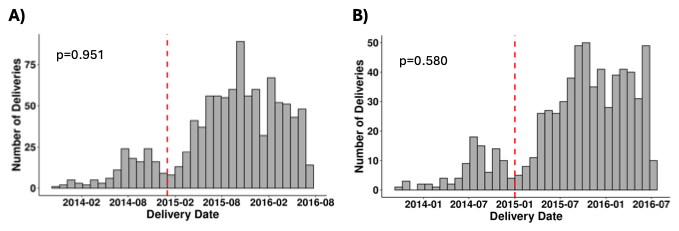

**Figure S2.** Flow diagram of data linkage and study sample

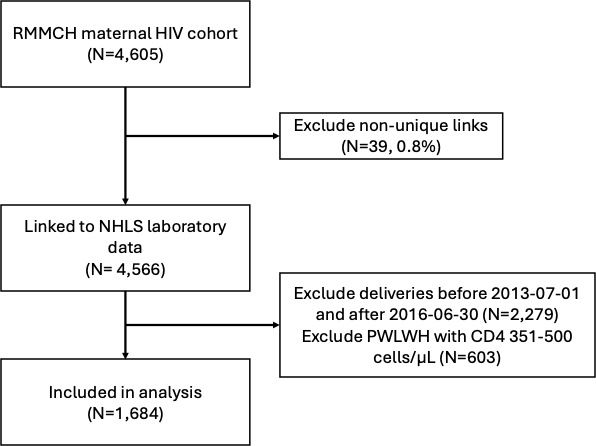

**Table S1.** Baseline characteristics at the threshold of Option B+ implementation: A) For the target population (CD4 >500); B) For the control population (CD4≤350)

A)

| Baseline measure |  | Below cutoff (Option B) | Above cutoff (Option B+) | Difference | P-value |
| --- | --- | --- | --- | --- | --- |
| Maternal age at delivery | Mean  (95% CI) | 27.3  (24.8, 29.7) | 30.8  (29.5, 32.1) | 3.5  (0.7, 6.3) | 0.013 |
| Newly on ART | %  (95% CI) | 71%  (54%, 88%) | 55%  (43%, 67%) | -15%  (-36%, 6%) | 0.151 |
| CD4 count | Mean  (95% CI) | 659  (591, 727) | 733  (677, 789) | 74  (-14, 162) | 0.102 |

B)

| Baseline measure |  | Below cutoff (Option B) | Above cutoff (Option B+) | Difference | P-value |
| --- | --- | --- | --- | --- | --- |
| Maternal age at delivery | Mean  (95% CI) | 31.0  (29.3, 32.7) | 30.2  (29.1, 31.2) | -0.8  (-2.8, 1.2) | 0.413 |
| Newly on ART | %  (95% CI) | 84%  (73%, 94%) | 83%  (76%, 90%) | -1%  (-13%, 12%) | 0.911 |
| CD4 count | Mean  (95% CI) | 222  (201, 244) | 210  (196, 224) | -13  (-44, 19) | 0.429 |

**Table S2.** Causal effect of Option B+ on postpartum retention, excluding deliveries from January to March 2015 (N=1,620)

| **A. Difference in Differences (DID)** | *Averages during each era* | | DID estimate (crude) | | DID estimate (adjusted)* | |
| --- | --- | --- | --- | --- | --- | --- |
|  | Option B | Option B+ | Pre/Post Difference | P-value | Pre/Post Difference | P-value |
| Target pop: CD4>500 | 42%  (32%, 52%) | 67%  (63%, 71%) | 19%  (5%, 32%) | 0.009 | 14%  (0%, 27%) | 0.049 |
| Control pop: CD4≤350 | 58%  (50%, 66%) | 64%  (61%, 68%) |  |  |  |  |
| **B. Regression Discontinuity (RD)** | *Predictions just before/after Option B+ policy change* | | RD estimate (crude) | | RD estimate (adjusted)* | |
|  | Just before Option B+ | Just after Option B+ | Pre/Post Difference | P-value | Pre/Post Difference | P-value |
| Target pop: CD4>500 | 41%  (22%, 61%) | 66%  (53%, 78%) | 24%  (1%, 48%) | 0.042 | 21%  (-1%, 42%) | 0.059 |
| Control pop: CD4≤350 | 56%  (41%, 72%) | 63%  (52%, 74%) | 7%  (-12%, 25%) | 0.494 | 9%  (-9%, 27%) | 0.349 |
|  | Difference in RD Estimates | | 18%  (-12%, 48%) | 0.248 | 12%  (-16%, 40%) | 0.396 |

**Estimates adjusted for maternal age at delivery (<26, 26-34, >34), ART history (newly on ART vs. continually on ART), and CD4 count value*

**Table S3.** Odds ratio of 6-24 months retention in care for ART-experienced birthing women (cases) vs. non-birthing women (controls)* (N=369)

| Characteristic | Odds ratio (95% CI) | P-value |
| --- | --- | --- |
| Birthing status (ref: Matched controls) |  |  |
| Cases | 1.68 (1.21, 2.34) | 0.002 |

**Matched by test type (VL/CD4), number of years in HIV care (0, 1, ≥2), district, age (*$\pm$*3 years), VL/CD4 test result (*$\pm$*25 units), and test date (*$\pm$*90 days). Each case was matched to 3 controls.*

**Table S4.** Comparison between Option B and Option B+ policies

|  | **Option A** | **Option B** | **Option B+** |
| --- | --- | --- | --- |
| **Date introduced in South Africa** | April 2010 | April 2013 | January 2015 |
| **Treatment guidelines** | CD4 ≤350: lifelong ART  CD4>350:  AZT (zidovudine) until delivery. At labor onset, single-dose nevirapine (sdNVP). At delivery, AZT+ lamivudine (3TC). After delivery, AZT+ lamivudine for seven days | CD4 ≤350: lifelong ART  CD4 >350: ART through postnatal period until one week after complete cessation of breastfeeding | Lifelong ART regardless of CD4 count |
| **Eligibility requirements** | Require CD4 test for eligibility | Require CD4 test for eligibility | Does not require CD4 test for immediate eligibility |
